## Supplementary Appendix 1 for "CATALYST trial protocol: A multicentre, open-label, phase II, multi-arm trial for an early and accelerated evaluation of the potential treatments for COVID-19 in hospitalised adults"

### Supplementary Appendix 1 – CATALYST investigators

The CATALYST investigators include the following;

Chief Investigator Prof. Tonny Veenith (University Hospitals Birmingham), deputy Chief Investigator Dr Benjamin Fisher (University of Birmingham), Dr Francis Mussai (University of Birmingham), Prof Gary Middleton (University of Birmingham), Dr Dhruv Parekh (University of Birmingham), Dr Anna Rowe (Cancer Research UK Clinical Trials Unit (CRCTU), University of Birmingham), Prof. Duncan Richards (University of Oxford), Dr Mathew Rowland (University of Oxford), Prof. Julian Bion (University of Birmingham), Daniel Slade (CRCTU, University of Birmingham), Prof. Simon Gates (CRCTU, University of Birmingham), Prof. Pam Kearns (CRCTU, University of Birmingham), Dr Rowena Sharpe (CRCTU, University of Birmingham), Dr Sarah Bowden (CRCTU, University of Birmingham), Prof. David Thickett (University Hospitals Birmingham), Prof. Julian Bion (University of Birmingham), Dr Tony Whitehouse (University Hospitals Birmingham), Dr James Scriven (University Hospitals Birmingham), Dr Mansoor Bangash (University Hospitals Birmingham), Prof. Fang Gao-Smith (University Hospitals Birmingham), Dr Jaimin Patel (University Hospitals Birmingham), Prof. Elizabeth Sapey (University Hospitals Birmingham), Prof. Mark Coles (University of Oxford), Prof. Peter Watkinson (University of Oxford), Prof. Naj Rahman (University of Oxford), Prof. Ling-Pei Ho (University of Oxford), Prof. Brian Angus (University of Oxford), Dr Alex Mentzer (University of Oxford), Dr Alex Novak (University of Oxford), Prof. Marc Feldmann (University of Oxford).

CRCTU staff: Charlotte Gaskell, Camilla Bathurst, Joseph van de Wiel, Alex Vince, Lili Evans, Rhian Jones, Karan Kaliri, Susie Mee, Karen James, Bushra Rahman, Karen Turner.

Trial Steering Committee: Prof. Michael Matthay (Chair), Prof. Danny McAuley (Deputy Chair), Prof. Paul Dark, Prof. Sir Andrew McMichael, Andrew Hall (Independent Statistician), Simon Farrell (PPI representative), Hannah Farrell (PPI representative)

Independent Data Management Committee: Prof. Adam Hill (Chair), Prof. Christina Yap (Independent Statistician), Prof. Anthony Gordon.

Scientific Advisory Board: Prof. Philip Newsome (Chair; Birmingham BRC Deputy Director, Clinician), Prof. Helen McShane (Deputy Chair, Oxford BRC Director, Clinician), Prof. Graham Cooke (Imperial College BRC Representative, Clinician), Prof. Duncan Richards (Director of Oxford CTRU, Clinician), Prof. Bryan Williams (UCL BRC Director, Clinician), Prof. Ling-Pei Ho (Translational Research Collaborative Representative, Clinician), Prof. Tonny Veenith (Chief Investigator, Clinician), Dr Ben Fisher (Deputy Chief Investigator, Clinician), Prof. Vincenzo Libri (CRF Representative, Clinician), Prof. Julian Bion (Clinical Trials Oversight Committee Representative, Clinician), Prof. Pamela Kearns (Trial Sponsor Representative, Director of CRCTU, Clinician), Dr Rowena Sharpe (Trial Management Group Representative), Prof. Simon Gates (Senior Biostatistician, CRCTU), Dr Rebecca Turner (Statistician).
