## Supplementary Appendix 2 for "CATALYST trial protocol: A multicentre, open-label, phase II, multi-arm trial for an early and accelerated evaluation of the potential treatments for COVID-19 in hospitalised adults"

### Supplementary Appendix 2 - Patient Information Sheets

<To be printed on hospital headed paper>

#### CATALYST - An early phase platform trial in (suspected) COVID-19

### Which treatment could lessen the severity of a (suspected or confirmed) COVID-19 infection when compared with usual care in an NHS setting?

#### Patient Information Sheet

##### Invitation to take part

CATALYST is a clinical trial being led by doctors and researchers based at the University of Birmingham and University Hospitals Birmingham NHS Foundation Trust, and in collaboration with the University of Oxford. You do not have to take part. Taking part is entirely voluntary, and you should only participate if you want to. Choosing not to take part in this trial will not affect your care in any way.

Before you decide whether you want to take part, it is essential for you to understand why the research is being done and what your participation will involve.

Please take time to read the following information carefully and ask your doctor if there is anything that is not clear or if you would like more information.

##### The purpose of the trial

There is currently no vaccine and few effective treatments for COVID-19. As COVID-19 is a new illness, we are constantly learning more about how it affects the human body. We know that the COVID-19 virus affects a number of different cells in your body, including a type of blood cell called a macrophage (immune cell), and that it can cause the number of these cells to increase in your body. To fight an infection, your immune cells produce proteins called cytokines and chemokines. These proteins can cause inflammation and at high levels can lead to damage in the tissues and organs in your body. Researchers believe this is why some people with COVID-19 infection become very ill.

Therapies we are testing:

A new, unlicensed drug called namilumab which has been tested in patients with arthritis and other inflammatory conditions. It may reduce inflammation in the body caused by the coronavirus. It is currently being given to patients with COVID-19 in a clinical trial in Italy. Namilumab is being provided free of charge by Izana Bioscience Limited (now part of Roivant Sciences) for use in this trial.

A drug called infliximab (Remsima) which is widely used to treat arthritis and other conditions may reduce inflammation in the body caused by the coronavirus. Infliximab is being provided free of charge by Celltrion Healthcare UK Limited for use in this trial.

Once this inflammation has been reduced, it may be possible that your immune system will adapt and fight off the virus more effectively.

Infliximab has not been tested in patients with COVID-19, and namilumab has only been given to a limited number of patients in Italy, therefore testing these drugs is the purpose of this trial. If any of the drugs benefit people with a (suspected or confirmed) COVID-19 infection in this trial, the drug will then be included in another larger-scale clinical trial being conducted throughout the UK, which is designed to compare treatments to find out which is the best at treating this infection.

This is a national trial, and some hospitals running it are unable to offer patients all of the therapies available within the trial. Your doctor will tell you which treatments are available at your hospital, and if applicable, which ones are not available.

##### Why have I been invited?

You have been invited to take part in this trial because you are aged 16 years or older, you have been admitted to hospital, and your doctor believes you have a COVID -19 infection and you have higher levels of C-reactive protein in your blood than normal (this increases when there's an infection or inflammation in your body).

##### What will happen to me if I take part?

- You will be agreeing to take part in a clinical trial and will be actively monitored for 28 days and longer if necessary.
- This clinical trial has three treatment groups or 'arms': one for each of the treatments listed above, and the other arm is 'usual care'. Usual care means that you will receive exactly the same treatment that you would receive in the hospital whether you decide to take part in the trial or not.
- The treatment you will receive is randomly allocated (chosen by chance) by a computer. Neither you nor your doctor will be able to choose the treatment you receive. Your research doctor or nurse will inform you of which treatment you will receive once they have entered you into the trial.
- If you are allocated the 'usual care' arm, you will receive the same medical care as all other patients being treated for a (suspected or confirmed) COVID-19 infection.

- If you are allocated to receive namilumab, you will receive this in addition to the usual medical care received by patients who have this disease. Namilumab will be given to you through a drip in a vein, usually in your arm, on one occasion only.
- If you are allocated to infliximab, you will receive this in addition to the usual medical care received by patients who have this disease. Infliximab will be given to you through a drip in a vein, usually in your arm, on one occasion only.
- You will have routine blood samples whilst you are in the hospital as part of 'usual care.' If you are allocated to the namilumab or infliximab arm, you may have more blood samples taken regularly before and during treatment to check it is safe for you to receive treatment.
- If you are a female of child bearing potential, a pregnancy test will be performed as part of the screening tests.
- We would also like your permission to collect samples for research. The collection of these samples is optional. This will only take place at certain hospitals. Your doctor will tell you if research samples are being collected at your hospital. This includes collecting samples of blood, and may include swabs from your nose and throat, and if you are placed on a ventilator, samples of the secretions from the tube placed in your windpipe to help you breathe may be taken. Samples will be collected on up to three separate occasions. These samples will be used in laboratory studies to gain a better understanding of COVID-19 and how patients respond to treatment.
- While you are in hospital your health and wellbeing will be monitored in accordance with usual care.

##### What are the potential benefits of taking part?

This research has been designed to help develop treatment for future patients with suspected or confirmed COVID-19 infection. It is important to understand that you may not directly benefit from taking part in this trial. The benefits for you as an individual are unknown.

##### What are the possible disadvantages and risks of taking part?

This trial is testing a new way of treating suspected or confirmed COVID-19 infection and you may have side effects from the treatment while taking part in the trial.

All of the treatments have been given to humans before but not in suspected or confirmed COVID-19 infection so there may be side effects that your doctors are not currently aware of, and these may be serious because of this an independent committee will be monitoring the safety of the treatments in this trial on a regular basis.

Everyone taking part in the trial will be monitored carefully for side effects. However, the doctors do not know all the side effects that may occur and we don't know how the drugs used in this trial will interact with the other drugs being used to treat (suspected or confirmed) COVID-19. Side effects may be mild or serious or may even be life-threatening. The doctors may give you medicines to help lessen side effects or the trial treatment may be postponed or stopped, depending on the side-effects they may experience.

Specific information is provided below for each of the trial treatments available in the trial is included below:

***Namilumab***

Namilumab is a new drug that is being tested to treat diseases that cause inflammation such as rheumatoid arthritis.

The side effects that are known so far are:

- Low neutrophil count (a type of white blood cell)
- Minor symptoms such as runny nose and headache
- Tachycardia (a fast heart beat)*

* A short change in heart rhythm was seen with no apparent harm in one person receiving namilumab in another trial.

***Infliximab***

Infliximab has been widely used for rheumatoid arthritis and other conditions for 20 years but it has some potential side effects. It has been used to treat patients with sepsis (serious infection) before on intensive care units and was demonstrated to be safe. The more common known side effects of infliximab when used to treat other conditions such as arthritis have been summarised below:

**While you are receiving Infliximab:**

<u>*Common side effects (experienced by 1 in 100 people) include:*</u>

- Allergic reaction: this can be mild but is sometimes severe and may even be life-threatening. For this reason you will be very carefully monitored while receiving the infliximab infusion and for the period afterwards
- Feeling and being sick
- Headache
- Flu-like symptoms

**During the days or weeks receiving Infliximab:**

<u>*Very common side effects (experienced by 1 in 10 people) include:*</u>

- Increased risk of infection. Infliximab can interfere with the body's ability to fight other infections caused by bacteria, fungi, other viruses such as hepatitis B, and tuberculosis. You will be monitored carefully for signs of infection other than COVID-19 and be offered appropriate treatment if another infection is detected.

*Common side effects (experienced by between 1 in 10 to 1 in 100 people) include:*

- Abnormal liver function tests. Blood tests will be done frequently to check that your liver is working properly.
- Some patients may experience worsening of psoriasis

***Pregnancy and Breast Feeding***

There is very little known about the effects of namilumab on an unborn baby and there is some information available about the infliximab on an unborn baby. As a precaution, women who are breast feeding are excluded from the trial and women of child bearing potential must have a negative pregnancy test prior to startng the trial. It is important that if you receive a trial treatment that you use adequate birth control if you (or your partner if you are male) are of child bearing potential. For namilumab, male and female participants should use effective contraception for 18 weeks after the last dose of drug. For infliximab, male and female participants should use contraception for 26 weeks after the last dose of drug.

Effective contraception is a method that has a failure rate of less than 1% a year when used correctly and all the time. Examples of these include:

- combined (oestrogen and progesterone containing) hormonal contraception e.g. the “pill”, patch
- progesterone only contraception (includes the “mini-pill”, injection, implant)
- Intrauterine device (IUD) or hormone-releasing system (IUS)
- vasectomised partner
- sexual abstinence

Recent guidelines in rheumatology patients suggests infliximab can be given to patients up until the 16$^{th}$ week of pregnancy. However, as this is the first time infliximab is being used in suspected or confirmed COVID-19 patients it is important that you understand the contraception information above.

It is also important that you do not breastfeed for 6 months after the last dose of namilumab or infliximab.

#### Is there any prohibited medication whilst I am on the trial?

Your trial doctor will look at the medicines that you are already taking and let you know whether you are able to still take them while you are taking part in the trial, or whether you would need to stop any of them.

##### What will happen if I don't want to carry on with the trial?

You can withdraw from the trial at any time without this having any effect on your medical care, however all information and blood samples already collected from your time on the trial will still be used.

##### Will my taking part in this trial be kept confidential?

All information collected about you for this trial will be subject to the General Data Protection Regulation 2018 and Data Protection Act 2018 for Health and Care Research and will be kept strictly confidential.

The University of Birmingham is the Sponsor for this trial. The University of Birmingham will be using information from your medical records in order to undertake this trial and will act as the data controller. This means that the University of Birmingham are responsible for looking after your information and using it properly. University of Birmingham and the NHS will keep identifiable information about you for at least 10 years after the trial has finished; this allows the results of the trial to be verified if needed.

In the Trial Office you will be identified by a unique trial number. In routine communication between your hospital and the Trial Office you will only be identified by trial number, initials and date of birth. Information about you, your health and wellbeing may be provided to the Trial Office on paper or electronically. We would also like to collect your NHS Number. This will allow researchers to collect information about your health and wellbeing from national records (e.g. Office for National Statistics, NHS Central Registries or other registries including those managed by NHS Digital) after the trial has ended. This will help us to determine the long-term impact of the trial treatment and COVID-19 on people's health.

By taking part in the trial you will be agreeing to allow research staff from the Trial Office to look at the trial records, including your medical records. It may be necessary to allow authorised personnel from government regulatory agencies (e.g. Medicines and Healthcare products Regulatory Agency (MHRA)), the Sponsor and/or NHS bodies to have access to information about you. This is to ensure that the trial is being conducted to the highest possible standards.

If you are randomised to receive namilumab or infliximab, pseudo-anonymised data from the trial may also be provided to the pharmaceutical company who is providing the drug you are given for safety monitoring or licensing purposes, where applicable. This is for your and others' protection to track the safety of the trial treatment. This may involve sending data outside of the United Kingdom to a European country of the United States of America. Your name and any identifying details (such as date of birth) will not be given to any of these parties.

We may be asked to share the trial information (data) we have collected with researchers running other studies, so that they can perform analysis on the data to answer other important questions about COVID-19.

These other researchers may be based in universities, NHS organisations, companies involved in health research and may be in this country or abroad. Non-identifiable summary information may also be shared with COVID-19 related UK government departments. Any such request is carefully considered by the trial researchers and will only be granted if the research is of high scientific standard and the necessary procedures and approvals are in place. The information will only be used for the purpose of health research and cannot be used to contact you or to affect your care. It will not be used to make decisions about future services available to you, such as insurance.

All individuals who have access to your information have a duty of confidentiality to you. Under no circumstances will you be identified in any way in any report, presentation or publication arising from this or any other research study.

You can withdraw your consent to our processing any more of your data at any time. Your rights to access change or move your information are limited, as we need to manage your information in specific ways in order for the research to be reliable and accurate. If you withdraw from the trial, we will keep the information about you and the samples that we have already obtained. Under the provisions of the General Data Protection Regulation 2018 and Data Protection Act 2018 you have the right to know what information the University of Birmingham has recorded about you. If you wish to view this information or find more about how we use this information, please contact Legal Services at Legal Services, University of Birmingham, Edgbaston, Birmingham, B15 2TT.

**What will happen to the samples I give?**

If you agree to take part in the optional research sample collection, the samples will be sent to laboratories based in the Universities of Birmingham or Oxford for storage and use in laboratory based research for the trial. If you give your consent, any samples leftover at the end of analysis for the trial, may be kept for future ethically approved research. These laboratory-based research projects will include genetic analysis of these samples. This DNA and RNA analysis is for scientific purposes and is not expected to provide findings of any clinical significance for you or your relatives, so the results will not be fed back to you. It is difficult to predict exactly what scientific developments there may be so we cannot give precise details of what research might be done.

No one using your samples for laboratory research will have access to your personal details. The samples sent to the University laboratories will be identified by your unique trial number only. All samples relating to you will be stored in accordance with the Human Tissue Act 2004.

##### What will happen to the results of the research trial?

At the end of the trial, the findings will be published in peer-reviewed medical and scientific journals. These publications will be available upon request from your trial doctor. We will also make a lay summary of the result available on the trial websites.

##### Who is organising and funding the research?

The trial is sponsored and being undertaken by the University of Birmingham in collaboration with University Hospitals Birmingham NHS Foundation Trust. The trial is being coordinated by the Cancer Research UK Clinical Trials Unit (CRCTU) within the University of Birmingham.

The trial is funded by an educational grant from UK Research and Innovation and drugs are being provided free of charge by the pharmaceutical companies.

##### Who has reviewed the trial?

All research in the NHS is looked at by an independent group of people called a Research Ethics Committee to protect your safety, rights, wellbeing and dignity. This trial has been reviewed and given favourable opinion by the East Midlands-Nottingham 2 Research Ethics Committee and by the NHS Health Research Authority. While the trial is ongoing the results will be reviewed regularly by an independent Data Monitoring Committee to ensure that it is appropriate to continue with the trial.

##### Expenses and Payments

As you will already be an inpatient in the hospital during the course of the trial you will not have to make any extra visits to participate in the trial and therefore will incur no additional expenses.

##### What if there is a problem?

If you have a concern about any aspect of this trial, you should ask to speak with your trial doctor who will do their best to answer your questions (see contact number at the end of this form). If you remain unhappy and wish to complain formally, you can do this through your hospital's Patient Advice and Liaison Services (PALS); they can be contacted by:

(Insert local contact details).

If you are harmed and this is due to someone's negligence then you may have grounds for legal action for compensation against the Sponsor of the trial (University of Birmingham) or the NHS Trust but you may have to pay your legal costs. NHS Trust Hospitals have a duty of care to all patients treated, whether or not the patient is taking part in a clinical trial, and the normal NHS complaints mechanisms will still be available to you (if appropriate).

#### Further information and contact details

Trial Doctor: ______________________________________________________

Research Nurse: ______________________________________________________

☎: __________________________ Emergency (24 hours) ☎: __________________________

<To be printed on hospital headed paper>

#### CATALYST - An early phase platform trial in (suspected or confirmed) COVID-19

### Which treatment could lessen the severity of a (suspected or confirmed) COVID-19 infection when compared with usual care in an NHS setting?

#### Personal Legal Representative Information Sheet

##### Invitation to take part

CATALYST is a clinical trial being led by the University of Birmingham and University Hospitals Birmingham NHS Foundation Trust in collaboration with the University of Oxford. You are being asked to assess the information contained within this information sheet on the patient's behalf, and make a decision based on the presumed wish of the patient on whether they would like to take part in the trial or not. The decision must be based on the patient's presumed will, and not based on your own views or objection.

Taking part is entirely voluntary, and choosing not to take part in this trial will not affect his/ her care in any way.

Before you decide whether the patient would want to take part, it is essential for you to understand why the research is being done and what his/her participation will involve. For this reason, the information contained within this personal legal representative information sheet is identical to that contained within the patient information sheet. The patient information sheet will be presented to the patient should he/ she clinically improve to the point of being able to assess the information to make an informed decision on whether to continue to take part in the trial or not.

Please take time to read the following information carefully.

##### The purpose of the trial

There is currently no vaccine and few effective treatments for COVID-19. As COVID-19 is a new illness, we are continually learning more about how it affects the human body. We know that the COVID-19 virus affects a number of different cells in the body, including a type of blood cell called a macrophage (immune cell), and that it can cause the number of these cells to increase in the body. To fight an infection, immune cells produce proteins called cytokines and chemokines. These proteins can cause inflammation and at high levels can lead to damage in the tissues and organs in the body. Researchers believe this is why some people with COVID-19 infection become very ill.

Therapies we are testing:
A new, unlicensed drug called namilumab which has been tested in patients with arthritis and other inflammatory conditions. It may reduce inflammation in the body caused by the coronavirus. It is currently being given to patients with COVID-19 in a clinical trial in Italy. Namilumab is being provided free of charge by Izana Bioscience Limited (now part of Roivant Sciences) for use in this trial.

A drug called infliximab (Remsima) which is widely used to treat arthritis and other conditions may reduce inflammation in the body caused by the coronavirus. Infliximab is being provided free of charge by Celltrion Healthcare UK Limited for use in this trial.

Once this inflammation has been reduced, it may be possible that the immune system will adapt and fight off the virus more effectively.

Infliximab has not been tested in patients with COVID-19, and nalimumab has only been given to a limited number of patients in Italy. Therefore, the purpose of this trial is to assess if these drugs may be helpful in treating COVID-19. If in this trial any drug shows an initial sign of benefit in (suspected or confirmed) COVID-19, then it will be included in another larger-scale clinical trial being conducted throughout the UK, which is designed to compare treatments to find out which is the best at treating this infection.

This is a national trial and some hospitals running this trial are unable to offer patients all of the therapies available within this trial. The doctor will tell you which treatments are available at the hospital, and if applicable, which ones are not available.

#### Why has my relative/ friend been invited?

Your relative/ friend has been invited to take part in this trial because they are aged 16 years or older, they have been admitted to hospital, and their doctor believes they have either a suspected or confirmed COVID -19 infection or have tested positive with a COVID-19 infection and they have higher levels of C-reactive protein in their blood than normal (this increases when there's an infection or inflammation in your body).

#### What will happen to my relative/ friend if they take part?

- You will agree for your relative/ friend to take part in a clinical trial, and they will be actively monitored for 28 days and longer if necessary.
- This clinical trial has three treatment groups or ‘arms’: one for each of the treatments listed above, and the other arm is ‘usual care’. Usual care means that they will receive the same treatment that everyone with a (suspected or confirmed) COVID-19 infection would get in the hospital whether you decide your relative/ friend takes part in the trial or not.
- The treatment they will receive is randomly allocated (chosen by chance) by a computer. Neither you nor your relative’s/ friend’s doctor will be able to select the treatment your relative/ friend receives. The research doctor or nurse will inform you of which treatment your relative/ friend will receive once they have entered them into the trial.
- If your relative/ friend is allocated the ‘usual care’ arm, they will receive the same medical care as all other patients being treated for a (suspected or confirmed) COVID-19 infection.
- If your relative/ friend is allocated to receive namilumab, they will get this in addition to the usual medical care received by patients who have this disease. Namilumab will be given to your relative/ friend through a drip in a vein, usually in their arm, on one occasion only.
- If your relative/ friend is allocated to receive infliximab, they will get this in addition to the usual medical care received by patients who have this disease. Infliximab will be given to your relative/ friend through a drip in a vein, usually in their arm, on one occasion only.
- Your relative/ friend will have routine blood samples while they are in the hospital as part of ‘usual care.’ If your relative/ friend is allocated to the namilumab or infliximab arm, they may have more blood samples taken regularly before and during treatment to check it is safe for them to receive treatment.
- If your relative/ friend are a female of childbearing potential, a pregnancy test will be performed as part of the screening tests.
- We would also like your permission to collect samples from your relative/ friend for research. The collection of these samples is optional. **This will only take place at certain hospitals**. The doctor will tell you if research samples are being collected at your relative/ friend’s hospital. This includes collecting samples of blood, and may include swabs from your relative/ friend’s nose and throat, and if they are placed on a ventilator, samples of the secretions from the tube placed in their windpipe to help them breathe may be taken. Samples will be collected on up to three separate occasions. These samples will be used in laboratory studies to gain a better understanding of COVID-19 and how patients respond to treatment.
- While your relative/ friend is in hospital, their health and wellbeing will be monitored in accordance with usual care.

##### What are the potential benefits of taking part?

This research has been designed to help develop a treatment for future patients with (suspected or confirmed) COVID-19 infection. It is important to understand that your relative/ friend may not directly benefit from taking part in this trial. The benefits for them as an individual are unknown.

##### What are the possible disadvantages and risks of taking part?

Your relative/ friend may have side effects from the treatment while taking part in the trial. This trial is testing a new way of treating (suspected or confirmed) COVID-19 infection.
All of the treatments have been given to humans before but not in (suspected or confirmed) COVID-19 infection, so there may be side effects that the doctors are not currently aware of, and these may be serious; because of this an independent committee will be monitoring the safety of the treatments in this trial regularly.

Everyone taking part in the trial will be monitored carefully for side effects. However, the doctors do not know all the side effects that may occur and we don't know how the drugs used in this trial will interact with the other drugs being used to treat (suspected or confirmed) COVID-19. Side effects may be mild or serious or may even be life-threatening. The doctors may give your relative/ friend medicines to help lessen side effects or the trial treatment may be postponed or stopped, depending on the side-effects they may experience.

Specific information is provided below for each of the trial treatments available in the trial is included below:

**Namilumab**

Namilumab is a new drug that is being tested to treat diseases that cause inflammation such as rheumatoid arthritis.
The side effects that are known so far are:

- Low neutrophil count (a type of white blood cell)
- Minor symptoms such as runny nose and headache
- Tachycardia (a fast heart beat)*

* A short change in heart rhythm was seen with no apparent harm in one person receiving namilumab in another trial.

**Infliximab**

Infliximab has been widely used for rheumatoid arthritis and other conditions for 20 years but it has some potential side effects. It has been used to treat patients with sepsis (serious infection) before on intensive care units and was demonstrated to be safe. The more common known side effects of infliximab when used to treat other conditions such as arthritis have been summarised below:

While your relative/ friend is receiving Infliximab:

- Common side effects (experienced by 1 in 100 people) include:
- Allergic reaction: this can be mild but is sometimes severe and may even be life-threatening. For this reason your relative/ friend will be very carefully monitored while receiving the iInfliximab infusion and for the period afterwards
- Feeling and being sick
- Headache
- Flu-like symptoms

**During the days or weeks receiving Infliximab:**

Very common side effects (experienced by 1 in 10 people) include:

- Increased risk of infection. Infliximab can interfere with the body's ability to fight other infections caused by bacteria, fungi, other viruses such as hepatitis B, and tuberculosis. Your relative/ friend will be monitored carefully for signs of infection other than COVID-19 and be offered appropriate treatment if another infection is detected.

Common side effects (experienced by between 1 in 10 to 1 in 100 people) include:

- Abnormal liver function tests. Blood tests will be done frequently to check that your relative/ friend's liver is working properly.
- Some patients may experience worsening of psoriasis

**Pregnancy and Breast Feeding**

There is very little known about the effects of namilumab on an unborn baby and there is some information available about the and infliximab on an unborn baby,. As a precaution, women who are breast feeding are excluded from the trial and women of child bearing potential must have a negative pregnancy test prior to startng the trial. It is important that if your relative/ friend receives a trial treatment that they use adequate birth control if they (or their partner if they are male) are of child bearing potential. For namilumab, male and female participants should use effective contraception for 18 weeks after the last dose of drug. For infliximab, male and female participants should use contraception for 26 weeks after the last dose of drug.

Effective contraception is a method that has a failure rate of less than 1% a year when used correctly and all the time. Examples of these include:

- combined (oestrogen and progesterone containing) hormonal contraception e.g. the "pill", patch
- progesterone only contraception (includes the "mini-pill", injection, implant)
- Intrauterine device (IUD) or hormone-releasing system (IUS)
- vasectomised partner

- sexual abstinence

Recent guidelines in rheumatology patients suggests infliximab can be given to female patients up until the 16th week of pregnancy. However, as this is the first time infliximab is being used in (suspected or confirmed) COVID-19 patients it is important your relative/ friend understands the contraception information above.
It is also important that any female patients do not breastfeed for 6 months after the last dose of namilumab or infliximab.

##### Is there any prohibited medication whilst my relative/ friend is on the trial?

Your relative/ friend's trial doctor will look at the medicines that they are already taking and let you/ your relative/ friend know whether they are able to still take them while they are taking part in the trial, or whether they would need to stop any of them.

##### What will happen if I don't want my relative/ friend to carry on with the trial?

You can withdraw your relative/ friend from the trial at any time without this having any effect on their medical care, however, all information and blood samples already collected from their time on the trial will still be used.

##### Will their taking part in this trial be kept confidential?

All information collected about your relative/ friend for this trial will be subject to the General Data Protection Regulation 2018 and Data Protection Act 2018 for Health and Care Research and will be kept strictly confidential.

The University of Birmingham is the Sponsor for this trial. The University of Birmingham will be using information from your relative/ friend's medical records in order to undertake this trial and will act as the data controller. This means that the University of Birmingham are responsible for looking after your relative/ friend's information and using it properly. The University of Birmingham and the NHS will keep identifiable information about them for at least ten years after the trial has finished; this allows the results of the trial to be verified if needed.

In the Trial Office, your relative/ friend will be identified by a unique trial number. In routine communication between the hospital and the Trial Office, your relative/ friend will only be identified by trial number, initials and date of birth. Information about your relative/ friend, their health and wellbeing may be provided to the Trial Office on paper or electronically. We would also like to collect your relative/ friend's NHS Number. This will allow researchers to collect information about their health and wellbeing from national records (e.g. Office for National Statistics, NHS Central Registries or other registries including those managed by NHS Digital) after the trial has ended. This will help us to determine the long-term impact of the trial treatment and COVID-19 on people's health.

By taking part in the trial, you will agree to allow research staff from the Trial Office to look at the trial records, including the medical records of your relative/ friend. It may be necessary to allow authorised personnel from government regulatory agencies (e.g. Medicines and Healthcare products Regulatory Agency (MHRA)), the Sponsor and/or NHS bodies to have access to information about your relative/ friend. This is to ensure that the trial is being conducted to the highest possible standards.

If the your relative/ friend is randomised to receive namilumab or infliximab, pseudo-anonymised data from the trial may also be provided to the pharmaceutical company who is providing the drug they are given for safety monitoring or licensing purposes, where applicable. This is for their and others' protection to track the safety of the trial treatment. This may involve sending data outside of the United Kingdom to a European country of the United States of America.  Your relative/ friend's name and any identifying details (such as date of birth) will not be given to any of these parties.

We may be asked to share the trial information (data) we have collected with researchers running other studies so that they can perform analysis on the data to answer other important questions about COVID-19. These other researchers may be based in universities, NHS organisations companies involved in health research and maybe in this country or abroad. Non-identifiable summary information may also be shared with COVID-19 related UK government departments.  Any such request is carefully considered by the trial researchers and will only be granted if the research is of high scientific standard and the necessary procedures and approvals are in place. The information will only be used for the purpose of health research and cannot be used to contact your relative/ friend or  affect their care. It will not be used to make decisions about future services available to them, such as insurance.

All individuals who have access to your relative/ friend's information have a duty of confidentiality to them. Under no circumstances will they be identified in any way in any report, presentation or publication arising from this or any other research study.

You can withdraw your consent to our processing any more of your relative/ friend's data at any time. Your rights to access change or move your relative/ friend's information are limited, as we need to manage their information in specific ways in order for the research to be reliable and accurate. If you withdraw your relative/ friend from the trial, we will keep the information about them and the samples that we have already obtained. Under the provisions of the General Data Protection Regulation 2018 and Data Protection Act 2018, your relative/ friend has the right to know what information the University of Birmingham has recorded about them.

If your relative/ friend wishes to view this information or find more about how we use this information, please contact Legal Services at Legal Services, University of Birmingham, Edgbaston, Birmingham, B15 2TT.

#### What will happen to the samples they give?

If you agree for your relative/ friend to take part in the optional research sample collection, the samples will be sent to laboratories based in the Universities of Birmingham or Oxford for storage and use in laboratory-based research for the trial. If you give your consent, any samples leftover at the end of analysis for the trial, may be kept for future ethically approved research. This laboratory-based research will include genetic analysis of these samples. This DNA and RNA analysis is for scientific purposes and is not expected to provide findings of any clinical significance for your relative/ friend or their relatives, so the results will not be fed back to your relative/ friend. It is difficult to predict precisely what scientific developments there may be so we cannot give precise details of what research might be done.

No one using your relative/ friend's samples for laboratory research will have access to their personal details. The samples sent to the University laboratories will be identified by your relative/ friend's unique trial number only. All samples relating to them will be stored in accordance with the Human Tissue Act 2004.

#### What will happen to the results of the research trial?

At the end of the trial, the findings will be published in in peer-reviewed medical and scientific journals. These publications will be available upon request from your relative/ friend's trial doctor. We will also make a lay summary of the result available on the trial websites.

#### Who is organising and funding the research?

The trial is sponsored and being undertaken by the University of Birmingham in collaboration with University Hospitals Birmingham NHS Foundation Trust . The trial is being coordinated by the Cancer Research UK Clinical Trials Unit (CRCTU) within the University of Birmingham.

The trial is funded by an educational grant from UK Research and Innovation and drugs are being provided free of charge by the pharmaceutical companies.

#### Who has reviewed the trial?

All research in the NHS is looked at by an independent group of people called a Research Ethics Committee to protect your relative/ friend's safety, rights, wellbeing and dignity. This trial has been reviewed and given favourable opinion by the East Midlands-Nottingham 2 Research Ethics Committee and by the NHS Health Research Authority. While the trial is ongoing the results will be reviewed regularly by an independent Data Monitoring Committee to ensure that it is appropriate to continue with the trial.

#### Expenses and Payments

As your relative/ friend will already be an inpatient in the hospital during the trial they will not have to make any extra visits to participate in the trial and therefore will incur no additional expenses.

#### What if there is a problem?

If you have a concern about any aspect of this trial, you should ask to speak with your relative/ friend‘s trial doctor who will do their best to answer your questions (see contact number at the end of this form). If you remain unhappy and wish to complain formally, you can do this through your relative/ friend‘s hospital’s Patient Advice and Liaison Services (PALS); they can be contacted by:

(Insert local contact details).

If your relative/ friend is harmed and this is due to someone’s negligence then your relative/ friend may have grounds for legal action for compensation against the Sponsor of the trial (University of Birmingham) or the NHS Trust but they may have to pay their legal costs. NHS Trust Hospitals have a duty of care to all patients treated, whether or not the patient is taking part in a clinical trial, and the normal NHS complaints mechanisms will still be available to your relative/ friend (if appropriate).

#### Further information and contact details

Trial Doctor: ______________________________________________________

Research Nurse: ______________________________________________________

☎: __________________________ Emergency (24 hours) ☎: __________________________

<To be printed on hospital headed paper>

### CATALYST - An early phase platform trial in (suspected or confirmed) COVID-19

#### Which treatment could lessen the severity of a (suspected or confirmed) COVID-19 infection when compared with usual care in an NHS setting?

#### Professional Legal Representative Information Sheet

##### Invitation to take part

CATALYST is a clinical trial being led by the University of Birmingham and University Hospitals Birmingham NHS Foundation Trustand in collaboration with the University of Oxford. You are being asked to consent on the basis of the presumed will of the patient by assessing the information contained within this information sheet. This applies both to the wish of the patient to take part, or to refuse to take part. Taking part is entirely voluntary, and choosing not to take part in this trial will not affect his/ her care in any way.

Before you decide whether the patient would want to take part, it is essential for you to understand why the research is being done and what his/her participation will involve. For this reason, the information contained within this professional legal information sheet is identical to that contained within the patient information sheet. The patient information sheet will be presented to the patient should he/ she clinically improve to the point of being able to assess the information in order to make an informed decision on whether to continue to take part in the trial or not.

Please take time to read the following information carefully.

##### The purpose of the trial

There is currently no vaccine and few effective treatments for COVID-19. As COVID-19 is a new illness, we are constantly learning more about how it affects the human body. We know that the COVID-19 virus affects several different cells in the body, including a type of blood cell called a macrophage (immune cell), and that it can cause the number of these cells to increase in the body. To fight an infection, immune cells produce proteins called cytokines and chemokines. These proteins can cause inflammation and at high levels can lead to damage in the tissues and organs in the body. Researchers believe this is why some people with COVID-19 infection become very ill.

Therapies we are testing:
A new, unlicensed drug called namilumab which has been tested in patients with arthritis and other inflammatory conditions. It may reduce inflammation in the body caused by the coronavirus. It is currently being given to patients with COVID-19 in a clinical trial in Italy. Namilumab is being provided free of charge by Izana Bioscience Limited (now part of Roivant Sciences) for use in this trial.

A drug called Infliximab (Remsima) which is widely used to treat arthritis and other conditions may reduce inflammation in the body caused by the coronavirus. Infliximab is being provided free of charge by Celltrion Healthcare UK Limited for use in this trial.

Once this inflammation has been reduced, it may be possible that the immune system will adapt and fight off the virus more effectively.

Infliximab has not been tested in patients with COVID-19, and namilumab has only been given to a limited number of patients in Italy. Therefore, the purpose of this trial is to assess if these drugs may be helpful in treating COVID-19. If any of the drugs benefit people with a (suspected or confirmed) COVID-19 infection in this trial, the drug will be included in another larger-scale clinical trial being conducted throughout the UK, which is designed to compare treatments to find out which is the best at treating this infection.

This is a national trial, and some hospitals running this trial are unable to offer patients all of the therapies available within this trial. The trial doctor will tell you which treatments are available at your hospital, and if applicable, which ones are not available.

##### Why has the patient been invited?

The patient has been invited to take part in this trial because they are aged 16 years or older, they have been admitted to hospital, and their doctor believes they have either a suspected or confirmed COVID -19 infection and the they have higher levels of C-reactive protein in their blood than normal (this increases when there's an infection or inflammation in your body).

##### What will happen to the patient if they take part?

- You will agree for the patient to take part in a clinical trial and they will be actively monitored for 28 days and longer if necessary.

- This clinical trial has three treatment groups or ‘arms’: one for each of the treatments listed above, and the other arm is ‘usual care’. Usual care means that the patient will receive exactly the same treatment that everyone with a (suspected or confirmed) COVID-19 infection would receive in the hospital whether you decide the patient takes part in the trial or not. The treatment the patient will receive is randomly allocated (chosen by chance) by a computer. Neither you nor the patient’s doctor will be able to choose the treatment they receive. The research doctor or nurse will inform you of which treatment the patient will receive once they have entered the patient into the trial.
- If the patient is allocated the ‘usual care’ arm, they will receive the same medical care as all other patients being treated for a (suspected or confirmed) COVID-19 infection.
- If the patient is allocated to receive namilumab, they will receive this in addition to the usual medical care received by patients who have this disease. Namilumab will be given to the patient through a drip in a vein, usually in their arm, on one occasion only.
- If the patient is allocated to receive infliximab, they will receive this in addition to the usual medical care received by patients who have this disease. Infliximab will be given to the patient through a drip in a vein, usually in their arm, on one occasion only.
- The patient will have routine blood samples whilst they are in the hospital as part of ‘usual care.’ If the patient is allocated to the namilumab or infliximab arm, the patient may have more blood samples taken regularly before and during treatment to check it is safe for them to receive treatment.
- If the patient is a female of childbearing potential, a pregnancy test will be performed as part of the screening tests.
- We would also like your permission to collect samples for research from the patient. The collection of these samples is optional. **This will only take place at certain hospitals.** The doctor will tell you if research samples are being collected at the patient’s hospital. This includes collecting samples of blood, and may include swabs from their nose and throat, and if they are placed on a ventilator, samples of the secretions from the tube placed in their windpipe to help them breathe may be taken. Samples will be collected on up to three separate occasions. These samples will be used in laboratory studies to gain a better understanding of COVID-19 and how patients respond to treatment.
- While the patient is in hospital their health and wellbeing will be monitored in accordance with usual care

#### What are the potential benefits of taking part?

This research has been designed to help develop a treatment for future patients with (suspected or confirmed) COVID-19 infection. It is important to understand that the patient may not directly benefit from taking part in this trial. The benefits for the patient as an individual are unknown.

#### What are the possible disadvantages and risks of taking part?

This trial is testing a new way of treating (suspected or confirmed) COVID-19 infection and the patient may have side effects from the treatment while taking part in the trial.

All of the treatments have been given to humans before but not in (suspected or confirmed) COVID-19 infection so there may be side effects that the doctors are not currently aware of, and these may be serious; because of this an independent committee will be monitoring the safety of the treatments in this trial on a regular basis.

Everyone taking part in the trial will be monitored carefully for side effects. However, the doctors do not know all the side effects that may occur and we don't know how the drugs used in this trial will interact with the other drugs being used to treat (suspected or confirmed) COVID-19. Side effects may be mild or serious or may even be life-threatening. The doctors may give the patient medicines to help lessen side effects or the trial treatment may be postponed or stopped, depending on the side-effects they may experience.

Specific information is provided below for each of the trial treatments available in the trial is included below:

**Namilumab**

Namilumab is a new drug that is being tested to treat diseases that cause inflammation such as rheumatoid arthritis.

The side effects that are known so far are:

- Low neutrophil count (a type of white blood cell)
- Minor symptoms such as runny nose and headache
- Tachycardia (a fast heart beat)*

* A short change in heart rhythm was seen with no apparent harm in one person receiving namilumab in another trial.

**Infliximab**

Infliximab has been widely used for rheumatoid arthritis and other conditions for 20 years but it has some potential side effects. It has been used to treat patients with sepsis (serious infection) before on intensive care units and was demonstrated to be safe. The more common known side effects of infliximab when used to treat other conditions such as arthritis have been summarised below:

Whilst receiving Infliximab:

- Common side effects (experienced by 1 in 100 people) include:

- Allergic reaction: this can be mild but is sometimes severe and may even be life-threatening. For this reason the patient will be very carefully monitored while receiving the iInfliximab infusion and for the period afterwards
- Feeling and being sick
- Headache
- Flu-like symptoms

During the days or weeks receiving Infliximab:

Very common side effects (experienced by 1 in 10 people) include:

- Increased risk of infection. Infliximab can interfere with the body's ability to fight other infections caused by bacteria, fungi, other viruses such as hepatitis B, and tuberculosis. The patient will be monitored carefully for signs of infection other than COVID-19 and be offered appropriate treatment if another infection is detected.

Common side effects (experienced by between 1 in 10 to 1 in 100 people) include:

- Abnormal liver function tests. Blood tests will be done frequently to check that the patient's liver is working properly.
- Some patients may experience worsening of psoriasis

**Pregnancy and Breast Feeding**

There is very little known about the effects of namilumab on an unborn baby and there is some information available about the and infliximab on an unborn baby.  As a precaution, women who are breast feeding are excluded from the trial and women of child bearing potential must have a negative pregnancy test prior to startng the trial.  It is important that if the patient receive's a trial treatment that they use adequate birth control if they (or their partner if they are male) are of child bearing potential.  For namilumab, male and female participants should use effective contraception for 18 weeks after the last dose of drug.  For infliximab, male and female participants should use contraception for 26 weeks after the last dose of drug.

Effective contraception is a method that has a failure rate of less than 1% a year when used correctly and all the time.  Examples of these include:

- combined (oestrogen and progesterone containing) hormonal contraception  e.g. the "pill", patch
- progesterone only contraception (includes the "mini-pill", injection, implant)
- Intrauterine device (IUD) or hormone-releasing system (IUS)
- vasectomised partner
- sexual abstinence

Recent guidelines in rheumatology patients suggests infliximab can be given to patients up until the 16th week of pregnancy. However, as this is the first time infliximab is being used in (suspected or confirmed) COVID-19 patients it is important that the patient understands the contracpetion information above.
It is also important that any female patients do not breastfeed for 6 months after the last dose of namilumab or infliximab.

##### Is there any prohibited medication whilst the patient is on the trial?

The trial doctor will look at the medicines that the patient is already taking and let you know whether the patient is able to still take them while they are taking part in the trial, or whether they would need to stop any of them.

##### What will happen if I don't want the patient to carry on with the trial?

You can withdraw the patient from the trial at any time without this having any effect on their medical care, however all information and blood samples already collected from the patient's time on the trial will still be used.

##### Will their taking part in this trial be kept confidential?

All information collected about the patient for this trial will be subject to the General Data Protection Regulation 2018 and Data Protection Act 2018 for Health and Care Research and will be kept strictly confidential.

The University of Birmingham is the Sponsor for this trial. The University of Birmingham will be using information from the patient's medical records in order to undertake this trial and will act as the data controller. This means that the University of Birmingham are responsible for looking after the patient's information and using it properly. The University of Birmingham and the NHS will keep identifiable information about the patient for at least 10 years after the trial has finished; this allows the results of the trial to be verified if needed.

In the Trial Office the patient will be identified by a unique trial number. In routine communication between their hospital and the Trial Office the patient will only be identified by trial number, initials and date of birth. Information about the patient, their health and wellbeing may be provided to the Trial Office on paper or electronically. We would also like to collect the patient's NHS Number. This will allow researchers to collect information about their health and wellbeing from national records (e.g. Office for National Statistics, NHS Central Registries or other registries including those managed by NHS Digital) after the trial has ended. This will help us to determine the long-term impact of the trial treatment and COVID-19 on people's health.

By taking part in the trial, you will be agreeing to allow research staff from the Trial Office to look at the trial records, including the patient's medical records. It may be necessary to allow authorised personnel from government regulatory agencies (e.g. Medicines and Healthcare products Regulatory Agency (MHRA)), the Sponsor and/or NHS bodies to have access to information about the patient. This is to ensure that the trial is being conducted to the highest possible standards.

If the patient is randomised to receive namilumab or infliximab, pseudo-anonymised data from the trial may also be provided to the pharmaceutical company who is providing the drug the patient is given for safety monitoring or licensing purposes, where applicable. This is for the patient's and others' protection to track the safety of the trial treatment. This may involve sending data outside of the United Kingdom to a European country of the United States of America. The patient's name and any identifying details (such as date of birth) will not be given to any of these parties.

We may be asked to share the trial information (data) we have collected with researchers running other studies, so that they can perform analysis on the data to answer other important questions about COVID-19. These other researchers may be based in universities, NHS organisations, companies involved in health research and may be in this country or abroad. Non-identifiable summary information may also be shared with COVID-19 related UK government departments. Any such request is carefully considered by the trial researchers and will only be granted if the research is of high scientific standard and the necessary procedures and approvals are in place. The information will only be used for the purpose of health research and cannot be used to contact you, the patient, or to affect the patient's care. It will not be used to make decisions about future services available to them, such as insurance.

All individuals who have access to the patient's information have a duty of confidentiality to them. Under no circumstances will the patient be identified in any way in any report, presentation or publication arising from this or any other research study.

You can withdraw your consent to our processing any more of the patient's data at any time. Your rights to access change or move the patient's information are limited, as we need to manage the information in specific ways in order for the research to be reliable and accurate. If you withdraw the patient from the trial, we will keep the information about them and the samples that we have already obtained. Under the provisions of the General Data Protection Regulation 2018 and Data Protection Act 2018 the patient has the right to know what information the University of Birmingham has recorded about the patient. If the patient wishes to view this information or find more about how we use this information, they should contact Legal Services at Legal Services, University of Birmingham, Edgbaston, Birmingham, B15 2TT.

#### What will happen to the samples the patient gives?

If you agree for the patient to take part in the optional research sample collection, the samples will be sent to laboratories based in the Universities of Birmingham or Oxford for storage and use in laboratory-based research for the trial. If you give your consent, any samples leftover at the end of analysis for the trial, may be kept for future ethically approved research. These laboratory-based research projects will include genetic analysis of these samples. This DNA and RNA analysis is for scientific purposes and is not expected to provide findings of any clinical significance for the patient or their relatives, so the results will not be fed back to the patient. It is difficult to predict precisely what scientific developments there may be so we cannot give precise details of what research might be done.

No one using the samples for laboratory research will have access to the patient's personal details. The samples sent to the University laboratories will be identified by the patient's unique trial number only. All samples relating to the patient will be stored in accordance with the Human Tissue Act 2004.

#### What will happen to the results of the research trial?

At the end of the trial, the findings will be published in peer-reviewed medical and scientific journals. These publications will be available to the patient upon request from their trial doctor. We will also make a lay summary of the result available on the trial websites.

#### Who is organising and funding the research?

The trial is sponsored and being undertaken by the University of Birmingham in collaboration with University Hospitals Birmingham NHS Foundation Trust The trial is being coordinated by the Cancer Research UK Clinical Trials Unit (CRCTU) within the University of Birmingham.

The trial is funded by an educational grant from UK Research and Innovation and drugs are being provided free of charge by the pharmaceutical companies.

#### Who has reviewed the trial?

All research in the NHS is looked at by an independent group of people called a Research Ethics Committee to protect the patient's safety, rights, wellbeing and dignity. This trial has been reviewed and given favourable opinion by the East Midlands-Nottingham 2 Research Ethics Committee and by the NHS Health Research Authority. While the trial is ongoing the results will be reviewed regularly by an independent Data Monitoring Committee to ensure that it is appropriate to continue with the trial.

#### Expenses and Payments

As the patient will already be an inpatient in the hospital during the course of the trial they will not have to make any extra visits to participate in the trial and therefore will incur no additional expenses.

#### What if there is a problem?

If you have a concern about any aspect of this trial, you should ask to speak with the patient's trial doctor who will do their best to answer your questions (see contact number at the end of this form). If you remain unhappy and wish to complain formally, you can do this through the hospital's Patient Advice and Liaison Services (PALS); they can be contacted by:

(Insert local contact details).

If the patient is harmed and this is due to someone's negligence then there may be grounds for legal action for compensation against the Sponsor of the trial (University of Birmingham) or the NHS Trust. NHS Trust Hospitals have a duty of care to all patients treated, whether or not the patient is taking part in a clinical trial, and the normal NHS complaints mechanisms will still be available to them (if appropriate).

#### Further information and contact details

Trial Doctor: ______________________________________________

Research Nurse: ______________________________________________

☎: ________________________ Emergency (24 hours) ☎: ________________________
