## Supplementary Appendix 3 for "CATALYST trial protocol: A multicentre, open-label, phase II, multi-arm trial for an early and accelerated evaluation of the potential treatments for COVID-19 in hospitalised adults"

#### Supplementary Appendix 3 - Informed Consent Forms

<To be printed on hospital headed paper>

### CATALYST- Early phase platform trial in (suspected or confirmed) COVID-19

#### Which treatment could lessen the severity of a (suspected or confirmed) COVID-19 infection when compared with usual care in an NHS setting?

#### Informed Consent Form

**Site:** ........................................................ **Patient's Trial Number:** ☐☐☐

**Investigator:** ........................................................ **EudraCT:** 2020-001684-89

**Please initial each box**

1. I confirm that I have read and understood the Patient Information Sheet (version ................... dated..........................) for the above trial. I have had the opportunity to consider the information, ask questions and have had these answered satisfactorily. ☐
2. I understand that my participation is voluntary and that I am free to withdraw at any time without giving any reason, without my medical care or legal rights being affected. ☐
3. I give permission for my initials, date of birth and NHS Number to be given to the Trial Office based within the University of Birmingham (Birmingham, UK) when I am randomised into the trial. ☐

Original to be kept in the Investigator Site File, 1 copy in hospital notes, 1 copy to the patient

4. I understand that relevant sections of my medical notes and data collected during the trial may be looked at by individuals from the Trial Office, regulatory authorities, the Sponsor (the University of Birmingham) and/or NHS bodies, where it is relevant to my taking part in this research. I give permission for these individuals to have access to my records.

5. I understand that data and samples from the trial may be provided to third parties (e.g. laboratories, pharmaceutical companies, other academic institutions or relevant UK government departments) for research, safety monitoring or licensing purposes, where applicable. I understand that this may involve sending data outside of the United Kingdom to a European country or the United States of America and that my name and any identifying details will NOT be given to these third parties, instead I will be identified by my unique Trial Number.

6. I understand that if I withdraw from the trial, any data collected up until the date of my withdrawal will be analysed and used as part of the research.

7. I agree to my GP being informed of my participation in this trial.

8. I agree to take part in the above trial.

**OPTIONAL**

9. I agree to the collection and storage of research samples for analysis as part of the CATALYST trial and for use in other ethically approved laboratory projects, which may include genetic analysis of these samples. I understand that I will not be personally informed of the results of these additional research projects and that if I withdraw from the trial any samples that have been collected up to the date of my withdrawal will not be destroyed.

| | | |
|---|---|---|
| ______________________________ | ____________________ | _________________________ |
| **Name of patient** | **Date** | **Signature** |
| ________________________________ | ____________________ | ____________________________ |
| **Name of person taking consent** | **Date** | **Signature** |

You must have signed the Site Signature & Delegation Log

Original to be kept in the Investigator Site File, 1 copy in hospital notes, 1 copy to the patient

<To be printed on hospital headed paper>

#### CATALYST- Early phase platform trial in (suspected or confirmed) COVID 19

#### Which treatment could lessen the severity of a (suspected or confirmed) COVID 19 infection when compared with usual care in an NHS setting?

#### Personal Representative Informed Consent Form

**Site:** ........................................................................ **Patient's Trial Number:** [ ][ ][ ]

**Investigator:** ........................................................................ **EudraCT:** 2020-001684-89

**Please initial each box**

1. I confirm that I been consulted about ……………………………………'s participant in this trial and have read and understand the Personal Legal Representative Information Sheet (version ................ dated........................) for the above trial and what it means to be a personal legal representative. I have had the opportunity to consider the information, ask questions and have had these answered satisfactorily. [ ]

2. I understand that his/her participation is voluntary and that I am free to change my opinion on what he/she would have wished for at any time without giving any reason, and without his/her medical care or legal rights being affected. [ ]

3. I understand that his/her direct informed consent will be obtained at the earliest opportunity. [ ]

4. I understand that his/her initials, date of birth and NHS Number will be given to the Trial Office based within the University of Birmingham (Birmingham, UK) when he/she is randomised to the trial.

5. I understand that relevant sections of his/her medical notes and data collected during the trial may be looked at by individuals from the Trial Office, regulatory authorities, the Sponsor (the University of Birmingham) and/or NHS bodies, where it is relevant to him/her taking part in this research.

6. I understand that data and samples from the trial may be provided to third parties (e.g. laboratories, pharmaceutical companies, other academic institutions or relevant UK government departments) for research, safety monitoring or licensing purposes, where applicable. I understand that this may involve sending data outside of the United Kingdom to a European country or the United States of America and that the patients name and any identifying details will NOT be given to these 3rd parties, instead he/she will be identified by his/ her unique Trial Number.

7. I understand that if he/she withdraws or is withdrawn from the trial, any samples and data that have been collected up to the date of withdrawal will be analysed and used as part of the research.

8. I agree to the patients GP being informed of his/ her participation in this trial.

9. In my opinion, the patient would have no objections to taking part in the above trial

**OPTIONAL**

10. I agree to the collection and storage of research samples for analysis as part of the CATALYST trial and for use in other ethically approved laboratory projects, which may include genetic analysis of these

samples. I understand that the patient will not be personally informed of the results of these additional research projects and that if he/ she withdraws from the trial any samples that have been collected up to the date of his/her withdrawal will not be destroyed.

**Name of patient:** ______________________________________________________________

_______________________________ ___________________ ___________________________

**Name of Personal Representative** **Date** **Signature**

_______________________________

**Relationship to patient**

_________________________________ ___________________ ___________________________

**Name of person taking consent** **Date** **Signature**

**You must have signed the Site Signature & Delegation Log**

<To be printed on hospital headed paper>

#### CATALYST- Early phase platform trial in (suspected or confirmed) COVID 19

### Which treatment could lessen the severity of a (suspected or confirmed) COVID 19 infection when compared with usual care in an NHS setting?

#### Professional Legal Representative Informed Consent Form

**Site:** ............................................................

**Patient's Trial Number:** | | | |

**Investigator:** ............................................................

**EudraCT:** 2020-001684-89

**Please initial each box**

1. I confirm that I been consulted about ……………………………………'s participant in this trial and have read and understand the Professional Legal Representative Information Sheet (version ................ dated........................) for the above trial and what it means to be a professional legal representative. I have had the opportunity to consider the information, ask questions and have had these answered satisfactorily. ☐

2. I understand that his/her participation is voluntary and that I am free to change my opinion on what he/she would have wished for at any time without giving any reason, and without his/her medical care or legal rights being affected. ☐

3. I understand that his/her direct informed consent will be obtained at the earliest opportunity. ☐

4. I understand that his/her initials, date of birth and NHS Number will be given to the Trial Office based within the University of Birmingham (Birmingham, UK) when he/she is randomised to the trial.

5. I understand that relevant sections of his/her medical notes and data collected during the trial may be looked at by individuals from the Trial Office, regulatory authorities, the Sponsor (the University of Birmingham) and/or NHS bodies, where it is relevant to him/her taking part in this research.

6. I understand that anonymised data and samples from the trial may be provided to third parties (e.g. laboratories, pharmaceutical companies, other academic institutions or relevant UK government departments) for research, safety monitoring or licensing purposes, where applicable. I understand that this may involve sending data outside of the United Kingdom to a European country or the United States of America and that the patients name and any identifying details will NOT be given to these third parties, instead he/ she will be identified by his/ her unique Trial Number.

7. I understand that if he/she withdraws or is withdrawn from the trial, any samples and data that have been collected up to the date of withdrawal will be analysed and used as part of the research.

8. I agree to the patients GP being informed of his/ her participation in this trial.

9. In my opinion, the patient would have no objections to taking part in the above trial

**OPTIONAL**

10. I agree to the collection and storage of research samples for analysis as part of the CATALYST trial and for use in other ethically approved laboratory projects, which may include genetic analysis of these

samples. I understand that the patient will not be personally informed of the results of these additional research projects and that if he/ she withdraws from the trial any samples that have been collected up to the date of his/her withdrawal will not be destroyed.

______________________________

**Name of patient:**

| ______________________________ | __________________ | __________________________ |
|---|---|---|
| **Name of Professional Legal Representative** | **Date** | **Signature** |

| ________________________________ | __________________ | __________________________ |
|---|---|---|
| **Name of person taking consent** | **Date** | **Signature** |

**You must have signed the Site Signature & Delegation Log**

Original to be kept in the Investigator Site File, 1 copy in hospital notes, 1 copy to the patient/ legal representative,
