## Supplementary Appendix 4 for "CATALYST trial protocol: A multicentre, open-label, phase II, multi-arm trial for an early and accelerated evaluation of the potential treatments for COVID-19 in hospitalised adults"

### Supplementary Appendix 4 – Statistical considerations

The simulations and tables below demonstrate the operating characteristics of a trial design with the chosen decision criteria, based on a simpler analysis of the area under the curve for sequential CRP data, with effect sizes informed from a dataset from 1026 hospitalised COVID-19 patients at Queen Elizabeth Hospital, Birmingham. In our simulations, we compared a traditional fixed trial design recruiting 120 patients with candidate adaptive designs. We present basic operating characteristics for the fixed design (Table 2A) and the chosen adaptive design (Table 2B). We studied six scenarios of treatment effect (see Table 5), and estimated, through simulation, the probability of a trial stopping early for "success" or "futility," and ultimately concluding success.

**Table 2A.** Operating characteristics for a fixed trial design of 120 patients.

| Scenario | Probability stopping early for success | Probability stopping early for futility | Overall probability of success | Mean number of patients |
|---|---|---|---|---|
| Null | 0 | 0 | 0.101 | 120 |
| A | 0 | 0 | 0.537 | 120 |
| B | 0 | 0 | 0.926 | 120 |
| C | 0 | 0 | 0.997 | 120 |
| D | 0 | 0 | 0.008 | 120 |
| E | 0 | 0 | 0 | 120 |

Scenarios A, B, and C are beneficial effects of the intervention with (true) treatment effects of 0.25, 0.5 and 0.75 standard deviations, "null" is zero treatment effect and D and E are harmful effects of 0.25 and 0.5 standard deviations. "success" and "futility" are defined as above.

**Table 2B.** Operating characteristics for an adaptive design with interim analyses at 40 and 80 patients.

| Scenario | Probability stopping early for success | Probability stopping early for futility | Overall probability of success | Mean number of patients |
|---|---|---|---|---|
| Null | 0.148 | 0.624 | 0.176 | 66 |
| A | 0.455 | 0.281 | 0.559 | 70 |
| B | 0.798 | 0.089 | 0.890 | 59 |
| C | 0.965 | 0.012 | 0.985 | 48 |
| D | 0.03 | 0.901 | 0.031 | 52 |
| E | 0.003 | 0.986 | 0.003 | 43 |

The adaptive design achieves similar probabilities of success in scenarios where the treatment effect is truly beneficial (A, B and C), and increases the probability of success only slightly if the intervention is harmful (D and E). There is some increase in the probability of success if the treatment effect is zero (Type I error) but this is offset by the very substantial reductions in the numbers of patients needed in all scenarios. Moreover, Type I error is not a serious problem as all interventions would be evaluated further in phase III trials.
