## Supplementary Appendix 5 for "CATALYST trial protocol: A multicentre, open-label, phase II, multi-arm trial for an early and accelerated evaluation of the potential treatments for COVID-19 in hospitalised adults"

### Supplementary Appendix 5 – Adverse event definitions

#### Adverse Event

Any untoward medical occurrence in a patient or clinical trial subject administered a medicinal product and which does not necessarily have a causal relationship with this treatment.

Comment:

An AE can therefore be any unfavourable and unintended sign (including abnormal laboratory findings), symptom or disease temporally associated with the use of an investigational medicinal product, whether or not related to the investigational medicinal product.

#### Adverse Reaction

All untoward and unintended responses to an IMP related to any dose administered.

Comment:

An AE judged by either the reporting Investigator or Sponsor as having causal relationship to the IMP qualifies as an AR. The expression reasonable causal relationship means to convey in general that there is evidence or argument to suggest a causal relationship.

#### Serious Adverse Event

Any untoward medical occurrence or effect that at any dose:

Results in death

Is life threatening*

Requires hospitalisation** or prolongation of existing inpatients' hospitalisation

Results in persistent or significant disability or incapacity

Is a congenital anomaly/birth defect

Or is otherwise considered medically significant by the investigator ***

Comments:

The term severe is often used to describe the intensity (severity) of a specific event. This is not the same as serious, which is based on patients/event outcome or action criteria.

* Life threatening in the definition of an SAE refers to an event in which the patient was at risk of death at the time of the event; it does not refer to an event that hypothetically might have caused death if it were more severe.

** hospitalisation is defined as an unplanned, formal inpatient admission, even if the hospitalisation is a precautionary measure for continued observation. Thus, hospitalisation for protocol treatment (e.g. line insertion), elective procedures (unless brought forward because of worsening symptoms) or for social reasons (e.g. respite care) are not regarded as an SAE.

*** Medical judgment should be exercised in deciding whether an AE is serious in other situations. Important AEs that are not immediately life threatening or do not result in death or hospitalisation but may jeopardise the subject or may require intervention to prevent one of the other outcomes listed in the definition above, should be considered serious.

#### Serious Adverse Reaction

An Adverse Reaction which also meets the definition of a Serious Adverse Event.

#### Suspected Unexpected Serious Adverse Reaction

A SAR that is unexpected i.e. the nature, or severity of the event is not consistent with the applicable product information.

A SUSAR should meet the definition of an AR, UAR and SAR.

#### Unexpected Adverse Reaction

An AR, the nature or severity of which is not consistent with the applicable product information (e.g. Investigator Brochure for an unapproved IMP or (compendium of) Summary of Product Characteristics (SPC) for a licensed product).

When the outcome of an AR is not consistent with the applicable product information the AR should be considered unexpected.
